# The COVID University Challenge: a Hazard Analysis of Critical Control Points Assessment of the Return of Students to Higher Education Establishments

**DOI:** 10.1101/2020.12.11.20247676

**Authors:** Kelly L. Edmunds, Laura Bowater, Julii Brainard, Jean-Charles de Coriolis, Iain Lake, Rimsha R. Malik, Lorraine Newark, Neil Ward, Kay Yeoman, Paul R. Hunter

## Abstract

The COVID-19 pandemic disrupted economies and societies throughout the world in 2020. Education was especially affected, with schools and universities widely closed for long periods. People under 25 years have the lowest risk of severe disease but their activities can be key to persistent ongoing community transmission. A challenge arose for how to provide education, including university level, without the activities of students increasing wider community SARS-CoV-2 infections.

We used a Hazard Analysis of Critical Control Points (HACCP) framework to assess the risks associated with university student activity and recommend how to mitigate these risks. This tool appealed because it relies on multi-agency collaboration and interdisciplinary expertise and yet is low cost, allowing rapid generation of evidence-based recommendations.

We identified key critical control points associated with university student’ activities, lifestyle and interaction patterns both on-and-off campus. Unacceptable contact thresholds and the most up-to-date guidance were used to identify levels of risk for potential SARS-CoV-2 transmission, as well as recommendations based on existing research and emerging evidence for strategies that can reduce the risks of transmission. Employing the preventative measures we suggest can reduce the risks of SARS-CoV-2 transmission among and from university students. Reduction of infectious disease transmission in this demographic will reduce overall community transmission, lower demands on health services and reduce risk of harm to clinically vulnerable individuals while allowing vital education activity to continue. HACCP assessment proved a flexible tool for risk analysis in a specific setting in response to an emerging infectious disease threat.

## 1. INTRODUCTION

Emerging infectious diseases present substantial threats to global health^1^. The current outbreak of SARS-CoV-2 that causes COVID-19 disease caught countries unaware and presented unprecedented challenges. To date the pandemic is linked to over 57.8 million reported cases and 1.3 million deaths (as at 22/11/20; WHO, 2020a). The long-term socio-economic harms of the pandemic are forecast with alarming numbers across multiple sectors including employment, global GDP, national debt and public service provision (World Bank, 2020). It is people under 25 years that are least in danger from the virus clinically (Russell et al., 2020), but this group arguably will shoulder the greatest long-term burden that result from for the current efforts to control the virus in-order to reduce societal harms. These dis-benefits/disadvantages include reduced education, reduced employment opportunities, living longer with degraded quality public services that in turn includes education that is likely to be highly resource-restricted in the near future (Maher, Hoang, & Hindery, 2020).

Early analyses suggests that closures of educational facilities had helped to reduce the outbreak in individual countries (Davies et al., 2020; Hunter et al., 2020; Li et al., 2020), through a variety of mechanisms. Although it remains unclear exactly how likely young children are to be infected by or transmit SARS-CoV-2, educational establishments contain adults that are interacting, collaborating and mixing. Also, young children are typically escorted to schools by adults who encounter other adults en route. Secondary and tertiary students often travel to school using public transport interacting highly with both each other and other adults en route and on site.

The prospect of shutting down educational facilities for long durations during the pandemic was not viable because the resultant unacceptable harm to both individuals and the wider economy was highly likely. For the age range 18-24 there was an additional consideration: a University College London survey found that loneliness during early lockdown was common in the age demographic (Leavey, Eastaugh, & Kane, 2020), an age when counts of social contacts normally peak rather than contract (Mossong et al., 2008). Full time home-study could increase loneliness and was often physically hampered by practical problems: many young adults live in rented accommodation without adequate quiet and dedicated space at home to study effectively (Leavey, Eastaugh, & Kane, 2020). Good internet access from home, or access to a PC/laptop, could also not be assured for all even in high income countries (Auxier & Anderson, 2020; Wakefield 2020). For individuals, a decision to suspend education/training and enter the labour market temporarily was not appealing as unemployment rates rose sharply (CTV News, 2020; Ergocun, 2020; Heath, 2020). For those students starting the new academic year in September 2020, approximately a third would have been enrolling in their first year of university study, which would bring with it additional unknowns.

A societal problem with the prospect of suspending university education for an uncertain but long duration was the medium-long term effects on professional training. Most high income countries recruit each year large numbers of newly trained graduates into a wide range of key worker professions, including nursing, medicine, allied health professions, social care, engineering, science and teaching. Suspending university education would increase existing shortages of essential degree-qualified workers across multiple sectors (Garcia & Weiss, 2019; Gilroy, 2019). Replacement recruitment of professionals from other parts of the world was unlikely given long training times required and pandemic disruption of education everywhere. It was also apparent that some universities faced severe financial crisis if students did not return to conventional tertiary education provision environment (Drayton & Waltmann, 2020); contraction of the tertiary education sector is not desirable as pandemic conditions demonstrated the value of degree trained professionals for response and infectious disease control.

Ensuring the continuation of tertiary education during the pandemic was imperative. Some degree courses can be effectively delivered in a tele-learning (online) format, but many courses still depend on practical work in laboratories, using university equipment as well as placement opportunities in external facilities that deliver in-person training. This reliance on specific facilities and types of training is especially true for vocational courses such as medicine, nursing, midwifery, physiotherapy is a challenge faced by our own institution which normally graduates hundreds of qualified health professionals each year. We were also aware that many non-health care students in our community, studying entirely online university degrees, lived on or near to campus and might still rely on other types of community locations (field work) or university facilities (such as the library or laboratories).

Therefore, the aim of this research was to identify the critical control points among the usual and necessary activities of university students – points where the risks of transmission were greatest but could also be mitigated and reduced –to make recommendations for mitigating these risks. The approach for this research involved the use of the Hazard Analysis of Critical Control Points (HACCP) framework; a low-cost tool for rapid assessment, that is simple for stakeholders to engage in, encourages multi-agency collaboration and interdisciplinary expertise whilst generating evidence-based recommendations (Edmunds et al., 2013). HACCP assessments originated in risk management of food production systems, but have been increasingly used to assess risks related to an emerging health threats and infectious diseases (Edmunds et al., 2013; Edmunds et al., 2016).

We applied the HACCP approach to the activities of university students and propose this method as a way to generate recommendations when dealing with a rapidly evolving infectious disease outbreak. We identify behaviours and practices linked to typical student activities which are likely to present risks for the transmission, both direct and indirect, of SARS-CoV-2. HACCP assessments are potentially powerful methods that can be used as rapid response tools in specific settings during emerging infectious disease outbreaks, as a precursor to more time-consuming and costly quantitative data collection, biomedical testing and clinical studies in specific settings.

## 2. METHODS

To complete our assessment of the risks of SARS-CoV-2 transmission during the return of students to UK university settings during the COVID-19 pandemic, we followed the first three HACCP principles (as used in previous studies into EIDs; Edmunds et al., 2013; Edmunds et al., 2016). The first stage of the process involved a series of virtual workshops and meetings amongst the HACCP team; all of whom are based at the higher education setting of the University of East Anglia (UEA) in Norwich, UK; a campus-based university with a student population of 17,429 in 2019/20 (of which 8,067 (46.3%) were new starters) (UEA 2020). The HACCP team includes experts in the fields of microbiology, infectious diseases, EIDs, public health, environmental health, science communication, student safety and student administrative services. The first workshop and subsequent follow-up discussions led to the creation of early versions of six flowcharts which consider different scenarios for student interactions on campus. These were i) on-campus teaching; ii) off-campus teaching; iii) living on campus; iv) living off-campus; v) non-teaching on-campus activities and vi) social activities. Following further consultation, the flowcharts were then critically analysed by the team and finalised, simplified versions produced. A hazard was considered to be a process that could take place within a higher education university setting that would provide the opportunity, at an unacceptable level of risk, for the transmission of SARS-CoV-2 to another person. Taking into account the likelihood of a situation occurring and the likelihood of SARS-CoV-2 transmission being able to occur within that situation, the hazards were then grouped into high, medium and low-risk categories. We generated recommendations for high-risk critical control points. The focus on high-risk CCPs is because our key objective was to make a set of recommendations that would do most to reduce transmission of SARS CoV-2 without being too onerous and likely to be followed. In concurrent circumstances (late 2020), an eradication strategy was not feasible in the UK. However, effective suppression of wide transmission to strongly limit harms from COVID-19 was stated in government policy and supported by most institutions and public experts (BBC, 2020; UK Government, 2020a). Therefore, in this HACCP exercise, we focused our recommendations on only situations which generate the majority of transmissions (“high risk”), but this HACCP exercise did not address situations where likelihood of transmission was low or medium. By focusing on high risk contexts, we sought to provide recommendations that would have maximum effect in keeping both the university and wider community prevalence of the infections at relatively low and thus manageable levels.

Following validation of the flowchart, the same team of experts focused on all of the high risk hazards and determined appropriate Critical Control Points (CCPs). A CCP is a point at which there is the opportunity to control, prevent or eliminate the risks for pathogen transmission. Critical limits were then set for each of the CCPs identified. These critical limits are thresholds used as preventative measures to control the hazards within the system. The outputs of each of these three principles were cross-referenced with the existing literature on SARS-CoV-2 epidemiology, prevention and control.

## 3. RESULTS

### 3.1 Hazard Analysis

Our HACCP assessment of the risks of SARS-CoV-2 transmission for students in UK university settings during the COVID-19 pandemic identified high-risk potentially hazardous practices linked to social interactions, teaching sessions and day-to-day on-campus activities (Table 1). Primary concerns were shared use of social spaces such as sports changing rooms, prayer facilities, study facilities including teaching rooms and rooms for the examination of patients for those students on medical and health science courses. When considering the sharing of student residences (both on and off campus), all those ordinarily living within the student residence were considered to come from the same household. Thus, social distancing and other protective measures such as the use of face coverings during interactions between them was not deemed necessary, unless one or more members of the household were presenting with symptoms of COVID-19 or had received a positive test result in the previous 14 days. All other activities and practices linked to students at higher education establishments were deemed to present a medium or low level of risk for the transmission of SARS CoV-2.

### 3.2 Critical Control Points and Critical Limits

In total, 22 high-risk CCPs were identified by this HACCP assessment. Each CCP is a point at which there is an opportunity to adopt measures to reduce the risks of transmission of SARS CoV-2 during the return of students to UK university settings. Following extensive consultation and cross-referencing with the existing literature, critical limits were then proposed for each high-risk CCP. In many cases, multiple recommendations were made for each CCP (Table 1).

Several of the critical limits suggested include fundamental infection control practices for coronaviruses, namely the correct and proper use of face coverings, employing appropriate hand hygiene measures and adopting appropriate social distancing measures, such as concurrently recommended physical distancing in the UK at a minimum of 1m+.

## 4 DISCUSSION

The HACCP framework allows for a rapid identification of the risks associated to a specific setting and evolving disease outbreak, in this case SARS-CoV-2 transmission to, from and between university students. The COVID-19 pandemic has highly disrupted education at all levels. It seems clear that shutting educational facilities can reduce community prevalence of SARS-CoV-2 but the harms long-term closure would cause to individuals and wider society are unacceptable. Therefore, strategies need to be found that enable most education delivery to continue, including forms of training that require physical use of education facilities.

No degree of pandemic preparedness can ever be fully comprehensive. This is because the nature of microbial contagions is that they tend to have unique features specific to the pathogenicity of the disease. The case of SARs-CoV-2 infection is distinctive because it is mostly harmless to a key demographic (young people) while COVID-19. However, there are many aspects of effective COVID-19 control that require tremendous disruptions to the lives of young people. Our HACCP assessment used a structured methodology to evaluate opportunities for mitigating risks of SARS-CoV-2 transmission associated with usual activities of university students, in an effort to facilitate their continued and essential education and training.

It seems clear that a proportion of SARS-CoV-2 transmission is from infected persons who lack symptoms: either pre-symptomatic and probably completely asymptomatic infections (Gandhi, Yokoe, & Havlir, 2020). The diversity of symptoms experienced is also large: COVID-19 is not simply an influenza-like illness with fever and cough as diagnostic symptoms (Sahoo, Jhunjhunwala, & Jolly, 2020). This situation presents unique challenges in terms of identifying cases and preventing them from passing the virus on. In peak prevalence periods, infection control relies on assuming that oneself and all other persons are potentially infected and therefore the only way to limit spread is through dramatically decreased contact with all other persons – even those with no contact or recent symptom history (Menon, 2020).

Our results feature recurring mitigation measures due to understanding of common disease transmission pathways and dynamics. Physical distancing from others (at least 2 metres) is believed to be especially effective because the predominant transmission pathway is via saliva droplets and possibly aerosols (WHO, 2020b). There is mixed quality evidence about the utility of wearing masks or face coverings to prevent COVID-19 in individuals (Brainard et al., 2020). In our own systematic review we concluded that wearing masks had some benefit for reducing transmission both when an infected person wore the mask and when a currently uninfected person wears one. However, we did not consider that mask wearing provided a guarantee of protection and was certainly not an alternative to other social distancing measures. The overall benefits of requiring populations to wear face coverings on wider community prevalence and subsequent mortality (Lyu & Wehby, 2020) is still not absolutely clear (Hunter et al., 2020).

High ventilation rates and outdoor settings are believed to be linked to lower risk of transmission (Bhagat et al., 2020). Good hand hygiene is basic infection control practice, as are measures to reduce the chances of fomite transmission (by sanitising any potentially shared objects). How to follow these principles in avoiding infection depends somewhat on setting and activity, such as meetings, religious worship, returns from family visits, queuing, interactions with household members, etc. We acknowledge in Table 1 that guidance and understanding of best infection control practices regarding COVID-19 are still emerging and recommend that students consult local guidance as it is published. Each transmission opportunity can be minimised or even eliminated through careful implementation of the critical limits outlined in Table 1.

At our own university, the response to COVID-19 has had three main components, covering communications, social distancing and testing and monitoring. Regular communications were sent to all students about COVID-19 safety measures and the University’s expectations around responsible behaviour. These were reinforced by enhanced security and monitoring of students on-campus, in accommodation and in other social settings, to discourage and prevent social gatherings that span households. Hand sanitiser and associated signage provided in teaching rooms and at key points in communal areas helped maintain a visible reminder of COVID-19 risk. To help ensure adequate distancing, one-way systems for navigating corridors within buildings and the narrower pathways outside on campus were introduced. In addition, a precautionary approach to 2m distancing in teaching rooms was pursued, based on shoulder-to-shoulder measurement and assuming some potential movement of torsos while sitting at desks (teaching room capacities were between 10 and 20% of normal).

At the start of autumn term, the University introduced an asymptomatic testing scheme in partnership with the Earlham Institute, a genomics research laboratory also located on the Norwich Research Park (Berger Gillam et al., 2020). The ‘Norwich Testing Initiative’ had capacity to analyse 500 PCR tests per day. All members of the university community (students and staff) who regularly visited campus or lived there were encouraged to get tested. Positive case students and their households were isolated for 10 days with non-symptomatic, non-positive housemates isolating for 14 days, with regular welfare support for those living off campus. There was daily monitoring of case numbers and patterns by University Safety Services and members of the University’s Executive Team, through a daily Major Operations Group meeting, to review trends and patterns and adjust the University’s response as necessary.

After students arrived back at UEA at the start of autumn term, the number of actively positive student cases being managed peaked at just 69 after 3 weeks, but fell to below 25 by week 6. There were just 2 cases that tested positive in the previous 10 days, by 9^th^ December. Of 360 cases in total detected over the first 11-weeks of monitoring, just three were members of staff involved in teaching. This suggests that the transmission reduction measures taken to prevent spread in teaching spaces were successful.

## 5 CONCLUSIONS

The COVID pandemic has delivered many epidemiological lessons and revealed numerous shortcomings in both local and international responses to emerging infectious diseases (EID). In the early stages of responding to an EID outbreak, the use of a response tool such as the HACCP framework allows for the rapid identification of risks and hazardous behaviours. Better risk reduction strategies can be generated after implementation of in-depth epidemiological studies, but such studies can take months or years to complete and produce results and recommendations. In contrast, the HACCP framework can be implemented fairly quickly using diverse informants and stakeholders. The long term consequences of the emergence of COVID-19 are proving difficult to predict, but we can still do much to limit pandemic harms. There is considerable merit in rapid responses methods to minimise harms linked to future disease outbreaks.

## Data Availability

Please see supplemental table.

## Acknowledgements

Professors Hunter and Lake as well as Dr. Brainard to the National Institute for Health Research Health Protection Research Unit (NIHR HPRU) in Emergency Preparedness and Response at King’s College London in partnership with Public Health England (PHE) and the University of East Anglia. The views expressed are those of the authors and not necessarily those of the NHS, the NIHR, UEA, the Department of Health or Public Health England. The authors are grateful to Jim Hunter of the University of East Anglia Safety Services staff for his valuable input and thank all those who have provided comment at any stage of the HACCP assessment or on this manuscript. As this study did not directly involve human subjects or data about individuals, ethical approval was not required.

---

**Supplementary Table 1.** Summary of the high risk activities identified through a Hazard Analysis of Critical Control Point (HACCP) assessment for the return of students to an on-campus university setting during the COVID-19 pandemic. This HACCP assessment considers the transmission of SARS-CoV-2 between individuals as the hazard under investigation.

| **Critical Control Point** | **Level of concern (high, medium, low)** | **Proposed Critical Limits and Recommendations** |
| --- | --- | --- |
| **1. Preventing arrival of virus in to the student household** | High | Ensure well-ventilated environment when interacting with individuals from outside the student household, outdoor interactions are lower risk than indoors (Qian et al., 2020; WHO, 2020c).<br>Wash hands with soap and water for at least 20 seconds after returning to student household. |
| **2. Shared car/minibus journey (including shared transport to placements, sporting events, field courses, social events etc.)** | High | Sit as far from others as possible, particularly avoiding sitting directly face to face with another passenger (UK Government, 2020b).<br>Open windows to increase ventilation (CDC, 2020a; GHHIN, 2020; Morawska & Milton, 2020; Qian et al., 2020; UK Government, 2020c, WHO, 2020c).<br>Wear face coverings in accordance with current national and university policy and guidance (WHO, 2020d).<br>Minimise journey time and where possible, keep journeys under 15 minutes in duration (ECDC, 2020; UK Government, 2020c). |
| **3. Returning to university household after time (overnight) away** | High | Self-isolate if had contact with confirmed case or told to self-isolate by public health officials or those working for their Test and Trace programmes (UK Government, 2020d).<br>Follow local guidance for the wearing of face coverings (WHO, 2020d).<br>Ensure good hand hygiene and washing practices (CDC, 2020b).<br>Maintain a minimum social distance of 2m as much as possible or at least 1m if other mitigations measures are in place, such as wearing face coverings (UK Government, 2020e). |
|  |  | Ensure well-ventilated environment (CDC, 2020a; GHIN, 2020; Morawska & Milton, 2020; Qian et al., 2020; UK Government, 2020c, WHO, 2020c).<br>Ensure clear guidance is available for students to follow. |
| **4. Interaction with people beyond university household (this includes the students having different placement groups to their other household members)** | High | Adhere to rules about group sizes (e.g. “Rule of six” implemented in September 2020) when socializing. Where possible minimize the number of university households involved in the interactions (Nielsen & Sneppen, 2020).<br>Minimise contact time between individuals and where possible keep interactions shorter than 15 minutes in duration (ECDC, 2020).<br>Ensure well-ventilated environment with windows open fully as much as possible (CDC, 2020a; GHIN, 2020; Morawska & Milton, 2020; Qian et al., 2020; UK Government, 2020c, WHO, 2020c).<br>Maintain a minimum social distance of 2m as much as possible or at least 1m if other mitigations measures are in place, such as wearing face coverings (UK Government, 2020e).<br>Wash hands with soap and water for at least 20 seconds after returning to student household. |
| **5. Sharing of personal items** | High –<br>Medium<br>depending on<br>what items are<br>being shared | Don’t share personal items! If unavoidable try to ensure a minimum of 72 hours between use by individuals from different households (van Doremalen, et al., 2020).<br>Sanitise items between uses by different individuals (CDC, 2020d; van Doremalen, et al., 2020).<br>Wash hands with soap and water for at least 20 seconds before and after exchange of items (CDC, 2020b; van Doremalen, et al., 2020). |
| **6. Indoor queueing /crowding** | High | Minimise queueing time by avoiding peak times. Keep queueing time to less than 15 minutes wherever possible (ECDC, 2020).<br>Observe and use space markers to help judge 2m distance (Nielsen & Sneppen, 2020; UK Government, 2020e); 2m is roughly 2 full adult arm lengths apart (Wikipedia, 2020). |
|  |  | Ensure well-ventilated environment (CDC, 2020a; GHIN, 2020; Morawska & Milton, 2020; Qian et al., 2020; UK Government, 2020c, WHO, 2020c).<br>Maintain a minimum social distance of 2m as much as possible or at least 1m if other mitigations measures are in place, such as wearing face coverings (UK Government, 2020e).<br>Wear face coverings in accordance with current national and university policy and guidance (WHO, 2020d).<br>Wash hands with soap and water for at least 20 seconds before and after queuing (CDC, 2020b; van Doremalen, et al., 2020). |
| **7. Sharing of study facilities** | High | Disinfect items between uses by different individuals (CDC, 2020c; van Doremalen, et al., 2020).<br>Ensure well-ventilated environment, open windows as much as possible (CDC, 2020a; GHIN, 2020; Morawska & Milton, 2020; Qian et al., 2020; UK Government, 2020c, WHO, 2020c).<br>Maintain a minimum social distance of 2m or at least 1m if other mitigation measures are in place, such as wearing face coverings (UK Government, 2020e).<br>Wear masks / face coverings in accordance with current national and university policy and guidance (WHO, 2020d) and avoid sitting face-to-face with another individual (UK Government, 2020b).<br>Wash hands with soap and water for at least 20 seconds before and after using study facilities (CDC, 2020b; van Doremalen, et al., 2020) and avoid touching your face (Macias et al., 2010). |
| **8. Outdoor queuing** | High – low depending on length of queue and time spent queuing | Minimise queueing time by avoiding peak times. Keep queueing time to less than 15 minutes when possible (ECDC, 2020).<br>Observe and use space markers to help you judge 2m distance coverings (UK Government, 2020e); 2m is roughly 2 full adult arm lengths apart (Wikipedia, 2020).<br>Maintain a minimum social distance of 2m as much as possible or at least 1m if other mitigations measures are in place, such as wearing face coverings (UK Government, 2020e; WHO, 2020d). |
|  |  | Wash hands with soap and water for at least 20 seconds before and after queuing (CDC, 2020b; van Doremalen, et al., 2020). |
| **9. Sharing of prayer facilities** | High | Adhere to rules about group sizes. Where possible minimize the number of university households involved in the interactions (Nielsen & Sneppen, 2020).<br>Minimise contact time between individuals, keeping the use of shared spaces to less than 15 minutes when possible (ECDC, 2020) and avoid singing or chanting and where deemed essential, limit the duration of singing (UK Government, 2020f).<br>Individuals to cleanse prior to attending prayer facility (UK Government, 2020f). If this is not possible, only one individual to use the cleansing room at a time with the room to be thoroughly cleaned and sanitized between uses (UK Government, 2020f; van Doremalen, et al., 2020).<br>Any items, such as a prayer mat, that are brought in to the facility must also be taken home immediately after use (UK Government, 2020f).<br>Ensure well-ventilated environment, with windows open fully where possible (CDC, 2020a; GHHIN, 2020; Morawska & Milton, 2020; Qian et al., 2020; UK Government, 2020c, WHO, 2020c).<br>Maintain a minimum social distance of 2m as much as possible or at least 1m if other mitigations measures are in place, such as wearing face coverings (UK Government, 2020e).<br>Wear face coverings in accordance with current national and university policy and guidance (WHO, 2020d).<br>Wash hands with soap and water for at least 20 seconds before and after using prayer facilities (CDC, 2020b; van Doremalen, et al., 2020). |
| **10. Sharing of changing facilities** | High | Adhere to rules about group sizes. Where possible minimize the number of university households involved in the interactions (Nielsen & Sneppen, 2020).<br>Minimise contact time between individuals, keeping the use of shared spaces to less than 15 minutes when possible (ECDC, 2020).<br>Do not share personal items. If unavoidable, sanitise shared items before using them (CDC, 2020d; van Doremalen, et al., 2020).<br>Ensure well-ventilated environment, with windows open fully when and where possible (CDC, 2020a; GHIN, 2020; Morawska & Milton, 2020; Qian et al., 2020; UK Government, 2020c, WHO, 2020c).<br>Maintain a minimum social distance of 2m as much as possible or at least 1m if other mitigations measures are in place, such as wearing face coverings (UK Government, 2020e).<br>Wear face coverings in accordance with current national and university policy and guidance (WHO, 2020d).<br>Wash hands with soap and water for at least 20 seconds before and after using shared facilities (CDC, 2020b; van Doremalen, et al., 2020). |
| **11. Sharing training/practice facilities** | High | Adhere to rules about group sizes. Where possible minimize the number of university households involved in the interactions (Nielsen & Sneppen, 2020).<br>Minimise contact time between individuals, keeping the use of shared spaces to less than 15 minutes when possible (ECDC, 2020) and avoid being face-to-face (UK Government, 2020b).<br>Do not share personal items. If sharing is unavoidable, disinfect items before using them (CDC, 2020d; van Doremalen, et al., 2020).<br>Ensure well-ventilated environment, with windows open fully where possible (CDC, 2020a; GHIN, 2020; Morawska & Milton, 2020; Qian et al., 2020; UK Government, 2020c, WHO, 2020c). |
|  |  | Maintain a minimum social distance of 2m as much as possible or at least 1m if other mitigations measures are in place, such as wearing face coverings (UK Government, 2020e).<br>Wash hands with soap and water for at least 20 seconds before and after using shared facilities (CDC, 2020b; van Doremalen, et al., 2020). |
| **12. Singing/Cheering** | High | Ensure well-ventilated environment, with windows open fully where possible (CDC, 2020a; GHIN, 2020; Morawska & Milton, 2020; Qian et al., 2020; UK Government, 2020c, WHO, 2020c).<br>Maintain a minimum social distance of 2m as much as possible or at least 1m if other mitigations measures are in place, such as wearing face coverings (UK Government, 2020e) and stand side-by-side rather than face-to-face (UK Government, 2020b).<br>Wear face coverings in accordance with current national and university policy and guidance (WHO, 2020d). |
| **13. Participation in conferences, meetings, etc.** | High | Where possible, make use of virtual meeting platforms.<br>Adhere to rules about group sizes when attending. Where possible minimize the number of university households involved in the interactions (Nielsen & Sneppen, 2020). .<br>Minimise contact time between individuals, keeping the use of shared spaces to less than 15 minutes when possible (ECDC, 2020).<br>Ensure well-ventilated environment, with widows open fully where possible (CDC, 2020a; GHIN, 2020; Morawska & Milton, 2020; Qian et al., 2020; UK Government, 2020c, WHO, 2020c).<br>Maintain a minimum social distance of 2m as much as possible or at least 1m if other mitigations measures are in place, such as wearing face coverings (UK Government, 2020e).<br>Wear face coverings in accordance with current national and university policy and guidance (WHO, 2020d). |
|  |  | Wash hands with soap and water for at least 20 seconds regularly throughout conference and before and after in-person meetings (CDC, 2020b; van Doremalen, et al., 2020). |
| **14. Sharing accommodation on field courses** | High | Avoid shared accommodation wherever possible. If unavoidable, adhere to rules about group sizes and where possible, minimize the number of university households involved in the interactions (Nielsen & Sneppen, 2020).<br>Do not share personal items. If sharing is unavoidable, sanitise any items before using them (CDC, 2020d; van Doremalen, et al., 2020).<br>Ensure well-ventilated environment, with windows open fully where possible (CDC, 2020a; GHHIN, 2020; Morawska & Milton, 2020; Qian et al., 2020; UK Government, 2020c, WHO, 2020c).<br>Maintain a minimum social distance of 2m as much as possible or at least 1m if other mitigations measures are in place, such as wearing face coverings (UK Government, 2020e).<br>Wear face coverings in accordance with current national and university policy and guidance (WHO, 2020d).<br>Wash hands with soap and water for at least 20 seconds before and after using shared facilities (CDC, 2020b; van Doremalen, et al., 2020).<br>Students must not attend field course if they have COVID symptoms<br>If student develops COVID symptoms during field course, isolation rules apply; consult current university and national policies |
| **15. Examining patients** | High-low depending on use of PPE | Examiner to wash hands before and after examining patient (CDC, 2020b; van Doremalen, et al., 2020).<br>Both examiner and patient to wear a mask and visor where possible (WHO, 2020d) and avoid being face-to-face (UK Government, 2020b).<br>Wear full PPE when examining the mouth, throat, and nose or when a patient has COVID symptoms (it is highly unlikely students will be allowed to examine such patients or do an Ear Nose Throat exam). |
| **16. Remaining within one room whilst on placement** | High | Ensure well-ventilated environment, with windows open fully where possible (CDC, 2020a; GHHIN, 2020; Morawska & Milton, 2020; Qian et al., 2020; UK Government, 2020c, WHO, 2020c).<br>Maintain a minimum social distance of 2m as much as possible or at least 1m if other mitigations measures are in place, such as wearing face coverings (UK Government, 2020e) and avoid being face-to-face (UK Government, 2020b).<br>Wear face coverings in accordance with current national and university policy and guidance (WHO, 2020d).<br>Students must not attend placement if they have COVID symptoms<br>Wash hands with soap and water for at least 20 seconds before and after using shared room (CDC, 2020b; van Doremalen, et al., 2020). |
| **17. Moving around within buildings** | High | Ensure good movement of people throughout shared spaces to minimize likelihood of crowding (Nielsen & Sneppen, 2020).<br>Minimise queueing/waiting time.<br>Follow any designated one-way systems and observe and use space markers to help judge 2m distance (UK Government, 2020e); 2m is roughly 2 full adult arm lengths apart (Wikipedia, 2020).<br>Ensure well-ventilated environment, with windows open fully where possible (CDC, 2020a; GHHIN, 2020; Morawska & Milton, 2020; Qian et al., 2020; UK Government, 2020c, WHO, 2020c).<br>Maintain a minimum social distance of 2m as much as possible or at least 1m if other mitigations measures are in place, such as wearing face coverings (UK Government, 2020e).<br>Wash hands with soap and water for at least 20 seconds before and after interactions with other individuals (CDC, 2020b). |
| **18. Sessions in teaching rooms** | High | Ensure well-ventilated environment, with windows open fully where possible (CDC, 2020a; GHHIN, 2020; Morawska & Milton, 2020; Qian et al., 2020; UK Government, 2020c, WHO, 2020c).<br>Ensure that rooms are assessed to ensure that a minimum social distance of 2m can be maintained as much as possible or at least 1m if other mitigations measures are in place, such as wearing face coverings (UK Government, 2020e).<br>Disinfect surfaces between teaching sessions (CDC, 2020d; van Doremalen, et al., 2020).<br>Wear face coverings in accordance with current national and university policy and guidance (WHO, 2020d).<br>Wash hands with soap and water for at least 20 seconds before and after using shared room (CDC, 2020b; van Doremalen, et al., 2020). |
| **19. Working in a group** | High | Where possible, make use of virtual meeting platforms.<br>Adhere to rules about group sizes. Where possible minimize the number of university households involved in the group working (Nielsen & Sneppen, 2020; UK Government, 2020e).<br>Minimise contact time between individuals, keeping the use of shared spaces to less than 15 minutes when possible (ECDC, 2020).<br>If possible, undertake group working outside. If not possible, ensure well-ventilated environment (CDC, 2020a; GHHIN, 2020; Morawska & Milton, 2020; Qian et al., 2020; UK Government, 2020c, WHO, 2020c). Disinfect surfaces between sessions (CDC, 2020d; van Doremalen, et al., 2020).<br>Maintain a minimum social distance of 2m as much as possible or at least 1m if other mitigations measures are in place, such as wearing face coverings (UK Government, 2020e).<br>Wash hands with soap and water for at least 20 seconds before and after group work or contact with another individual (CDC, 2020b; van Doremalen, et al., 2020). |
| **20. Seating in teaching rooms** | High | Maintain a minimum social distance of 2m as much as possible or at least 1m if other mitigations measures are in place, such as wearing face coverings (UK Government, 2020e).<br>Wear face coverings in accordance with current national and university policy and guidance (WHO, 2020d).<br>Wash hands with soap and water for at least 20 seconds before and after group work or contact with another individual (CDC, 2020b; van Doremalen, et al., 2020).<br>Disinfect surfaces between teaching sessions (CDC, 2020d; van Doremalen, et al., 2020). |
| **21. Undertaking physical exams such as OSCEs/ consultation skills / physical manipulations** | High-low depending on use of PPE | Ensure 2m distance between students after exam<br>Examiner, student, and actor (if used) to keep 2m distance (UK Government, 2020e), or wear a mask and visor if social distancing cannot be maintained (WHO, 2020d).<br>Ensure well-ventilated environment, with windows open fully where possible (CDC, 2020a; GHGIN, 2020; Morawska & Milton, 2020; Qian et al., 2020; UK Government, 2020c, WHO, 2020c).<br>Students to wash hands before entering and when leaving each station (CDC, 2020b; van Doremalen, et al., 2020).<br>Examiner to wipe down any surfaces touched by students before next student enters the station (van Doremalen, et al., 2020).<br>Avoid mouth-to-mouth in emergency scenarios and instead provide single-use bag valve masks<br>Students to bring their own equipment (e.g. stethoscope and pen) to each station (van Doremalen, et al., 2020).<br>Use models/dummies instead of actors/patients for examinations where possible; sanitise models/dummies between users (CDC, 2020d; van Doremalen, et al., 2020). |
| **22. Provision of ad hoc in-person support (e.g. advisors, last minute support etc.)** | High | If possible and appropriate, provide support outside or using video conferencing. If not possible, ensure support is given in a well-ventilated environment (CDC, 2020a; GHIN, 2020; Morawska & Milton, 2020; Qian et al., 2020; UK Government, 2020c, WHO, 2020c).<br>Adhere to rules about group sizes (Nielsen & Sneppen, 2020).<br>Minimise contact time between individuals, keeping the use of shared spaces to less than 15 minutes when possible (ECDC, 2020).<br>Maintain a minimum social distance of 2m as much as possible(UK Government, 2020e) or at least 1m if other mitigations measures are in place, such as wearing face coverings<br>Wash hands with soap and water for at least 20 seconds before and after in-person meetings (CDC, 2020b; van Doremalen, et al., 2020). |

